# Assessing the impact of a symptom-based mass screening and testing intervention during a novel infectious disease outbreak: The case of COVID-19

**DOI:** 10.1101/2020.02.20.20025973

**Authors:** Yang Ge, Brian Kenneth McKay, Shengzhi Sun, Feng Zhang, Andreas Handel

**Author notes:** Corresponding author: Yang Ge, Andreas Handel, Corresponding author.

## Abstract

A symptom-based mass screening and testing intervention (MSTI) can identify a large fraction of infected individuals during an infectious disease outbreak. China is currently using this strategy for the COVID-19 outbreak. However, MSTI might lead to increased transmission if not properly implemented. We investigate under which conditions MSTI is beneficial.

## Background

Rapid detection and isolation of infected individuals is important for containing infectious disease outbreaks (1). A symptom-based mass screening and testing intervention (MSTI), is an approach that can identify a significant fraction of infected individuals. In the current COVID-19 outbreak, several countries such as China, Italy and US are using the MSTI strategy to identify as many cases as possible. A problem for MSTI is that many novel pathogens show symptoms that are similar to common infections, e.g., general respiratory symptoms. This can lead to rapid overload of the health system (2). If testing for MSTI is performed in health facilities, such a situation might directly lead to a worsening of the outbreak if there is transmission risk associated with MSTI (3).

## Objective

We explore how the potential benefit of MSTI, taking into consideration MSTI associated transmission risk.

## Methods and Findings

We consider a scenario where a novel pathogen causes symptoms that are similar to circulating pathogens, e.g., general respiratory symptoms. At any given time during an outbreak, a proportion, *P*, of those showing symptoms will be infected with the novel pathogen. For symptomatic patients who are asked to stay home (H), the expected number of transmissions of the novel pathogen caused by that person is the average number of transmissions caused by someone who is infected with that pathogen (reproductive number, R), multiplied by the probability the person is infected with the new pathogen, *T*_*H*_ =*RP*. An alternative option is to implement MSTI. Mass screening tries to identify all individuals who show symptoms consistent with the novel pathogen. For individuals who go through an MSTI, there are two possible scenarios. If the symptomatic individual is infected with the novel pathogen (probability *P*), we assume that they are correctly identified and placed under isolation and treatment. Screened individuals are assumed to have reduced transmission risks, *R*_*I*_ *P*. If the health system is strained, those patients undergoing testing without being infected with the novel pathogen (probability1 − *P*) may become infected during MSTI. We denote this probability of becoming infected with *F*. Since these individuals test negative, we assume they will be sent home and have the same transmission risk as individuals asked to remain at home. Thus, the total transmission potential for a person undergoing MSTI (S) is *T*_*S*_= *R*_*I*_ *P + RF* (*1*− *P*). Similar expressions for an individual’s mortality risk under either scenario are shown in the SM.

We can define the transmission risk ratio (TRR) as *TRR* = *Ts* / *TH*. If *TRR* <1, MSTI reduces overall transmission. As a best-case scenario, we consider a situation where isolation following a positive test is perfect, *R*_*I*_ = 0, (other scenarios are explored in the SM). This gives *TRR =* (1− *P*) *F*/*P*. Figure 1 shows a heatmap for TRR (on a log scale) for different values of *F* and *P*. The solid line is given by *F* = *P/* (1− *P*), at which TRR=1 (thus their log is 0). This line marks the threshold at which MSTI changes from being beneficial to being harmful. While hard numbers for the quantities *F* and *P* are impossible to obtain, we used previously published data to obtain very rough estimates for *F* and *P* ranges for COVID-19, Ebola and Measles (see SM for details). Our estimates suggest that MSTI is clearly beneficial for Ebola, this is less certain for Measles and COVID-19.

**Figure:**
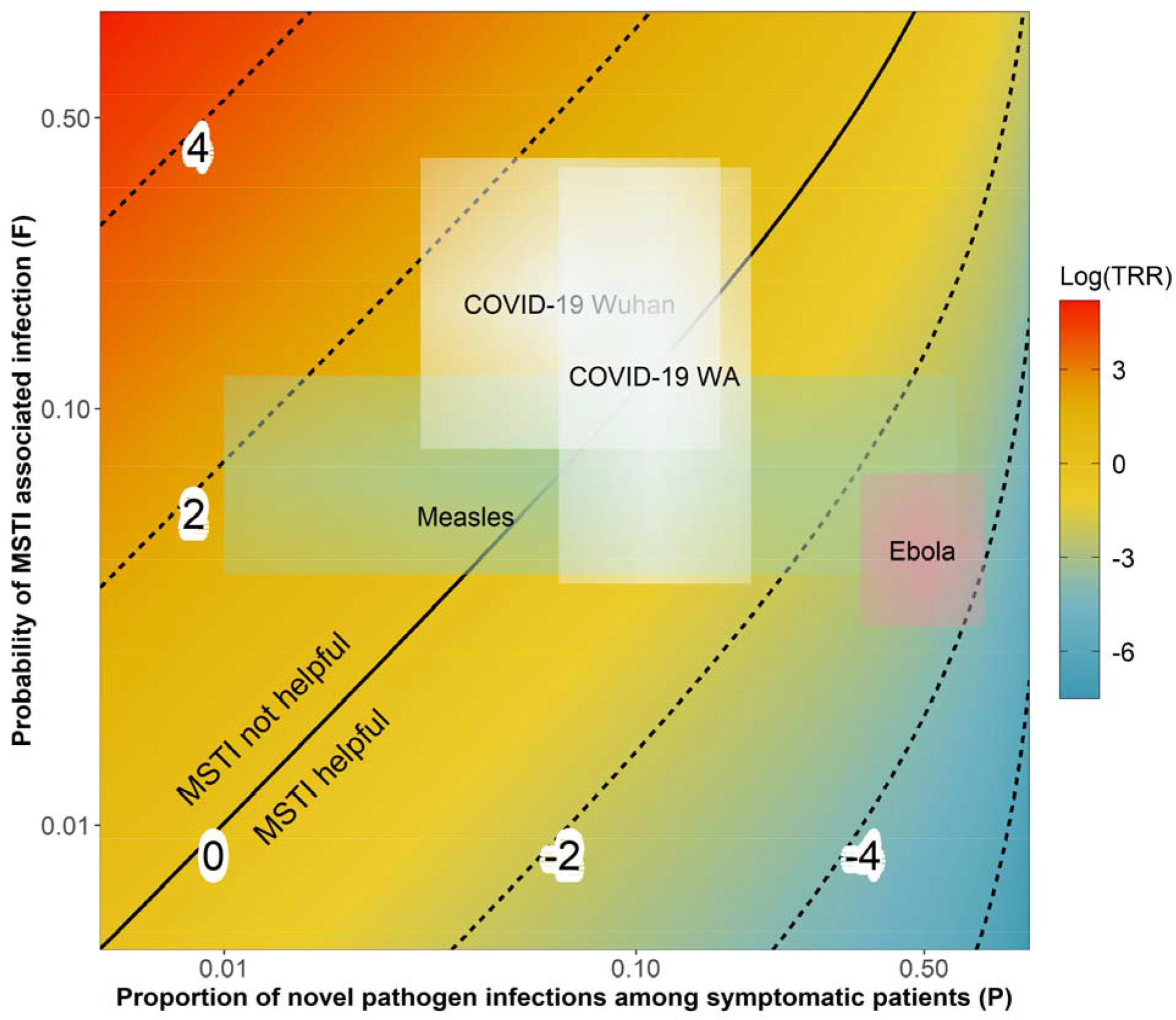
Transmission risk ratio (log scale) as a function of novel pathogen prevalence among individuals showing symptoms, *P*, and testing-site related infection risk factor, *F*. For TRR the threshold at which MSTI becomes beneficial is 1, thus for the log shown here this threshold is 0. Indicated in color are estimates for *P* and *F* ranges for 3 different pathogens (see SM for details).

## Discussion

For MSTI to be beneficial, one needs to minimize *F* and maximize *P* (4). An approach to reduce *F* is through testing sites separate from the usual healthcare facilities. *P* can be increased by having a more specific case definition (ideally without losing sensitivity). This will also reduce the total number of individuals going to testing sites, likely reducing *F*. Overall, our analysis suggests that MSTI can be useful if the probability of transmission at testing sites is less than the probability that a symptomatic person is infected with the novel pathogen. Both *F* and *P* will likely vary between settings and thus should be evaluated for a specific setting, e.g. a specific country, if MSTI is considered as a potential control strategy.

## Data Availability

We used previously published data.

## Notes

**Conflict of interest: None**

**Funding statement: None**

### Competing Interest Statement

The authors have declared no competing interest.

### Funding Statement

No fund supporting this research.

## References

1. Fraser C, Riley S, Anderson RM, Ferguson NM. Factors that make an infectious disease outbreak controllable. Proceedings of the National Academy of Sciences. 2004;101(16):6146–51.

2. Lipsitch M, Hayden FG, Cowling BJ, Leung GM. How to maintain surveillance for novel influenza a h1n1 when there are too many cases to count. The Lancet. 2009;374(9696):1209–11.

3. Chowell G, Abdirizak F, Lee S, Lee J, Jung E, Nishiura H, et al. Transmission characteristics of mers and sars in the healthcare setting: A comparative study. BMC medicine. 2015;13(1):210.

4. Kucharski AJ, Camacho A, Checchi F, Waldman R, Grais RF, Cabrol J-C, et al. Evaluation of the benefits and risks of introducing ebola community care centers, sierra leone. Emerging Infectious Diseases. 2015 Mar;21(3):393–9.

